# Social determinants of adult health outcomes in Sickle cell disease: a scoping review protocol

**DOI:** 10.1101/2023.08.15.23294030

**Authors:** Alphanso Blake, Nadia Bennett, Trevor Ferguson, Seeromanie Harding, Monika Asnani

**Affiliations:** Epidemiology Research Unit, Caribbean Institute for Health Research, The University of the West Indies, Mona, Jamaica; School of Life Course Sciences, Faculty of Life Sciences & Medicine, King’s College London, London, UK; Sickle Cell Unit, Caribbean Institute for Health Research, University of the West Indies, Mona, Jamaica

**Keywords:** Life course, social determinants, sickle cell disease, morbidity, scoping review protocol

## Abstract

**Objective:** To summarize the evidence and identify gaps in the literature related to the impact of social factors on health outcomes across the life course for persons with Sickle Cell Disease.

**Introduction:** Sickle cell disease (SCD) is a group of genetic diseases where abnormal hemoglobin is associated with recurrent complications across the life course which results in increased morbidity or death. The social determinants of health may impact health outcomes throughout the life course of these individuals. Better understanding of the effect that these factors and their complications have may influence management approaches both medically and socially to improve quality of life.

**Inclusion criteria:** All studies published in English between 1973 and 2023 of persons of all ages living with SCD (SS, SC, SB^0^ SB^+^), examining factors related to the social determinants of health will be included. Conference abstracts, editorials and opinion papers will be excluded.

**Methods:** Methods proposed by the Joanna Briggs Institute (JBI) and by Westphaln and colleagues in 2021 will be used. The electronic databases Academic Search Ultimate (EBSCO), Open Science Framework and PubMed will be searched using a pre-specified search strategy developed with stakeholder input. All citations will be reviewed by two independent reviewers with disagreements adjudicated by a third reviewer. The screening process will be reported in a Preferred Reporting Items for Systematic Reviews Extensions for Scoping reviews (PRISMA-ScR) flow diagram. The Rayyan software will be used to manage the review process.

## Introduction

Sickle cell disease (SCD) is one of the most common hereditary disorders globally, predominantly affecting populations of African and Indian descent (1, 2). Although the pathophysiological effects of the condition has been extensively studied, the non-genetic factors such as socioeconomic and climate factors has not been as widely reviewed, especially in low income settings (3). The USA has less than 1% of persons with SCD, in Populations in Africa the prevalence varies between 2 and 30% and, and in Jamaica 10% of the population carries the HbS gene (3-5). The four commonest types of SCD in Jamaica are homozygous SS, SC, SB^0^ thalassemia, and SB^+^ thalassemia. The disease is characterized by multiple acute and chronic sequele (6) which can have deleterious effects on an individuals’ development in childhood and social and economic positions in adulthood. These effects are a result of missed school days which are associated with lower academic ability and poorer job prospects. (6, 7).

### Social Determinants of Health

Factors such as access to healthcare, parental occupation, community socioeconomic status and educational attainment continue to play a significant role in the quality-of-life, and impact health outcomes. These factors can be grouped under a major heading called Social Determinants of Health (SDoH)(8). Social determinants of health have been defined as the conditions in which people are born, grow, work, live, age, and the wider set of forces and systems shaping the conditions of their daily life (9). There is a myriad of factors, of which the World Health Organization (WHO) has outlined ten, namely social gradient, stress, early life, social exclusion, work, unemployment, social support, addiction, food and transport as a lens through which social determinants of health can be classified or viewed (8). The above-mentioned factors can be used to assess an individual’s situation as they progress throughout their course of life. Adopting a life course approach to the review of the conditions people are exposed to rather than a snapshot approach can offer a more thorough and complete understanding of challenges faced at all ages and how the factors interact with each other(10).

The Sustainable Development Goals look at good health and wellbeing (Goal 3) and Reducing inequalities (Goal 10), the SDoH construct is a central ingredient to all action (10, 11). The SDoH construct does well to incorporate the work of George Engel 1977 which involves the conceptualization of the biopsychosocial model to care (12). Engels posits that in order to understand a person’s medical condition adequately, one needs to factor in their psychological and social situation which forms an interplay with their biological situation(12). Authors in recent times built on Engel’s work and emphasize that approaches to handling social inequities in health should consider the determinants which often lies outside the health system such as poverty, income, employment status and education. (10).

### Morbidity in SCD

Morbidity factors include acute and chronic severe bone pain may affect the extremities, chest and back (2). Acute pain is especially common in adolescents and young adults and is seen more frequently in persons with the homozygous HbSS genotype. Severe pain episodes are often managed at home without the involvement of health providers. Persons with the condition have at least 0.8 (≈ 1) significant episode of pain per year, with over 5% having 3 - 10 episodes yearly (1).

Acute chest syndrome is the second most common complication in SCD and is characterized by chest pain, fever, dyspnea, and decreased oxygen saturation (13). It is a form of acute lung injury which can occur after an acute pain event and carries a risk for respiratory and multiorgan failure(13). Researchers have found that it can result in significant mortality, of up to 9 or 10% of cases in adults with SCD (2, 13, The condition is associated with an average hospitalization of 10.5 days and decreased long-term survival and as such is considered a medical emergency treatable in hospital immediately (13).

Acute stroke is a serious and potentially debilitating complication in SCD; this includes ischaemic and haemorrhagic events and presents as a medical emergency(2). Ischaemic stroke is caused by an occlusion of a cerebral artery resulting from deoxygenated red blood cells which deform into a rigid sickle shape, it can also be a result of complications of other SCD-related events(2). Ischaemic stroke may be preceded by, transient ischaemic attacks, which are characterized by sudden onset of focal weakness or alteration in mental state, lasting less than twenty-four hours while ischaemic strokes last longer than 24 hours (2). Children with SCD have a 3 times greater risk of acute stroke than other children and by the age of 45 years, 25% of individuals with SCD would have had a stroke (2).

Sickle cell leg ulcers are a common complication and their prevalence is affected by the individual’s age, genotype, and geographical location (15). It is the most common cutaneous presentation of SCD and occurs as early as the teenage years (16, 17). The condition is most common in persons with HbSS and persons with West African origins, wherein Jamaica has a very high (70% - 80%) prevalence, while other areas report a lower prevalence (18). Additionally, a study from the USA has reported leg ulceration to be more common in males than in females(15). The variety of ulcers that have been found can be categorized into three groups, namely, single ulcers, recurrent ulcers and chronic recurrent ulcers (19). Chronic leg ulceration can result in intense, chronic and unrelenting pain which is difficult to manage (16). Leg ulcerations negatively impact one’s quality of life, resulting in depression in some cases (16). Regardless of the many complications of SCD, more patients with the condition are living longer due to the introduction of newborn screening, simple early childhood interventions, better health maintenance and the introduction of novel therapies (20). The median age of death, however, remains lower than the general population wherein individuals with SCD live up to 22 fewer years (54 vs 76 years) and have a quality-adjusted life expectancy of half that of individuals without SCD (33 vs 67 years) (7, 21). The median age of death in the Jamaican cohort is 42.3 years(22).

The life course approach to chronic diseases theorizes that the accumulation and or interaction of socio-economic, biological and psychosocial experiences throughout the life course of an individual will ultimately impact adult health outcomes, a number of models have been put forward to explain life course (23).

A preliminary search of the PubMed database was conducted as a pilot exercise. One Scoping review study was found which looked at the relationship between sickle cell disease and social determinants of health by Khan 2022 (24). The results of this scoping review study had fifty-nine articles which were classified under 5 SDoH headings, namely, Neighbourhood and built Environment (11 studies), Health and Health Care (15 studies), Social and Community Context (18 studies), Education (5 studies) and Economic Stability (11 studies). This list may not fit under all the WHO classification listed above except for economic stability (work) and social and community context (social support). Most of the studies described by Khan were from the United States (43 studies) and the United Kingdom (6 studies). Gaps detailed by the study include that most research was limited to children and caregivers, and none were found that assessed the longitudinal impact of social determinants of health (24). No grey literature was searched in Khan’s work, exclusion of grey literature may be considered a limitation as smaller studies presented at conferences, as part of academic work or that have not reached publication status would have been missed. No other systematic reviews or scoping reviews were found in the search.

This scoping review will aim to identify and map the social determinants that have shown associations with health outcomes including pain events, hospitalizations, acute chest syndrome, stroke, and quality of life across the life course of individuals with sickle cell disease. This study will also aim to ascertain the longitudinal association of SDoH factors and health outcomes in SCD to address the gaps found in the study by Khan (24). Finally, these findings will assist in filling knowledge gaps related to the area and provide a basis for future research and development.

### Review question(s)

What are the social determinants that have been shown to be associated with health outcomes across the life course of persons with Sickle Cell Disease?

I. What measures have been used for measuring SDoH in published studies on Sickle cell disease?
II. What is the existing evidence of the association between Social Determinants of Health (SDOH) and Sickle Cell Disease (SCD) over the life course?
III. How many studies have examined the intersectionality between Social Determinants of Health (SDOH) and Sickle Cell Disease (SCD)?

## Inclusion criteria

### Participants

This review will consider studies that have been reported on persons with SCD (HbSS, HbSC, Hb Sβ0 thal, Hb Sβ+ thal). All study designs will be included.

### Concept

This review will consider studies that explore predictors such as socioeconomic status, parental occupation, environmental factors, and health service accessibility. Secondly, outcomes will be assessed in manuscripts which included quality of life, hospitalization, complications (leg ulcers, acute chest syndrome, strokes, painful crises, kidney disease) and death.

### Context

This review will consider studies that were published between 1973 and 2023 (inclusive), from all world regions. Studies not published in English and opinion papers will be excluded.

### Types of sources

This scoping review will consider qualitative, quantitative, and mixed methods study designs for inclusion. Studies done using a qualitative methodology are expected to offer rich, thick data. Finally, systematic, and scoping reviews that match the inclusion criteria will be considered, and their reference list combed to identify additional studies that are relevant to the research question. Text, conference abstracts and opinion papers will not be considered, and neither will studies which are not written in English.

## Methods

The scoping review will be conducted following the JBI methodology for scoping reviews documented by Peters and colleagues in 2015 and supported by recommendations from Westphaln and colleagues in 2021 (20, 21). It will also be conducted in line with the Preferred Reporting Items for Systematic Reviews and Meta-Analyses extensions for Scoping Reviews (PRISMA-ScR (25-27).

### The Review team

The study team will consist of three reviewers, AB the primary researcher, NB who has experience in systematic review methodologies and MA who is on the team as a senior researcher and content expert.

Consultations will be sought from relevant stakeholders inclusive of specialist SCD physicians, social worker, nurse, patients with SCD and their proxies. This is detailed in the consultation section below.

### Search strategy

The search strategy will identify published and unpublished studies. Using an iterative process Academic Search Ultimate (EBSCO), Open Science Framework and PubMed databases will be searched. The PubMed database was searched on June 9^th^, 2023, as part of a piloting exercise. The search terms are detailed in appendix 1.

The reference list of systematic reviews identified as well as studies identified for inclusion will be screened for any potentially relevant articles which may have been missed by the search strategy. Grey literature sources will include American Society of Hematology, Sickle Cell in Focus and Global Congress on Sickle Cell Disease conference documents which were identified by AB and MA the content expert. All citations retrieved will be stored in Endnote 20 and uploaded into Rayyan. All duplicates will be removed.

### Study/Source of evidence selection

Following the search, all identified records will be collated and uploaded into EndNote 20 and duplicate removed. Following a pilot test, titles and abstracts will then be screened by two independent reviewers for assessment against the inclusion criteria for the review. Potentially relevant papers will be retrieved in full, and their citation details imported into the Raayan software for full assessment and review. The full text of selected citations will be assessed in detail against the inclusion criteria independently by AB and NB. Any disagreements on suitability for inclusion in the study will be arbitrated by MA who will have the final decision. Reasons for the exclusion of full-text papers that do not meet the inclusion criteria will be recorded and reported in the scoping review. The results of the search will be reported in full in the final scoping review and presented in a PRISMA flow diagram(25).

### Data extraction

Data will be extracted from papers included in the scoping review by AB and NB using a data extraction tool developed by NB. The data extraction form will capture the author’s name, study year, study location, community of residence, hospitalization, social amenities scores, educational status, parental educational status, and occupation as SDoH proxies as well as outcome variables (Stroke, acute chest, pain events, hospitalization, and mortality). Additionally, the population of the study, sample size, aims, methodology, study design used, key results and limitations will be extracted. Data abstraction will be conducted by AB and NB independently and consolidated to ensure the completeness of the information retrieved. MA will make the final decision on any questions raised or disagreements between AB and NB. Authors of papers will be contacted to request missing or additional data, where required. The data will thereafter be summarized and coded in a simple qualitative format in excel. A draft of the extraction form is available in Appendix II. In keeping with the iterative nature of a scoping review, the extraction form will be amended if necessary. All modifications made to the data extraction form will be detailed in the final report.

### Data analysis and presentation

The consultation team along with the review team will come together to examine the findings to ensure that they are relevant and in keeping with the proposed objectives/intentions of the study. Qualitative data will be sorted by two members of the research team. Results will be coded as themes and classified into morbidity factors and social determinants of health factors associated with each morbidity factor.

Quantitative data such as the number of participants in each study, age, and age of onset of morbidity factor will be summarized as means and standard deviation or median and interquartile range as appropriate. Following the feedback from all members of the consultation team, data will be summarized and represented in a tabular form to all members for consolidation of findings. Finally, a narrative summary will accompany each table, thus describing how the findings connect to the intended aim of the review.

## Consultation

Westphaln 2021 considers consultation an integral part of a scoping review and not just an optional feature (28). The consultation exercise is expected to add legitimacy to the findings and the trustworthiness of the consultants is expected to be transparent and rigorous(29). This Sickle Cell Unit (SCU) is the specialised comprehensive care facility for persons with SCD in Jamaica and hence provides a useful proxy for the experiences of persons on the island. A consultation team including members of staff from the SCU and SCD patient support group will be assembled. The team members will be a social worker who works closely with the population of persons with SCD, 2 specialist SCD doctors and a specialist SCD nurse who has over 10 years of experience with the population. Additionally, two patients, two parent/guardians of patients (not linked), and two members from SCD peer support groups will be included on the consultation team. These individuals will be recruited using a purposive sampling approach. The sessions will be recorded and transcribed for use during the period of the study and disposed of at the end. Consultation is key as it provides valuable feedback in which knowledge can be integrated which may not necessarily be accessible via academic literature (28). These may include cultural practices, historical context and newly developed approaches which may not have been published yet (28). The consultation team will provide input at the beginning and at the end of the review to ensure that appropriate terms are being used and that the research question stays true to the intended objective.

## Data Availability

All data produced in the present study are available upon reasonable request to the authors

## Dissemination and ethics

Ethical approval was obtained from the Mona Research Ethics Committee (MREC) at the University of the West Indies (# CREC-MN.0154, 2022/2023).

## Acknowledgements

Mrs. Faith McKoy-Johnson, a librarian, provided support with the development of the search strategy.

## Declarations

No special declarations are to be made.

## Author contributions

AB designed the protocol and wrote the manuscript. NB, TF, SH, and MA contributed to the protocol design and guided the writing of the manuscript. MA and TF contributed to content development and NB focused on design principles for a review of this nature. All authors agree with the final version of the manuscript for submission.

## Appendix I: Search strategy

#PubMed June 9^th^, 2023

| Search | Query | Records<br>retrieved |
| --- | --- | --- |
| #1 | ((("Anemia, Sickle Cell"[Mesh] OR "Anemias, Sickle Cell" [TW] OR "Sickle Cell Anemias" [TW] OR "Hemoglobin S Disease" [TW] OR "Disease, Hemoglobin S" [TW] OR "Hemoglobin S Diseases" [TW] OR "Sickle Cell Anemia" [TW] OR "Sickle Cell Disorders" [TW] OR "Sickle Cell Disorder" [TW] OR "HbS Disease" [TW] OR "Sickle Cell Disease" [TW] OR "Cell Disease, Sickle" [TW] OR "Cell Diseases, Sickle" [TW] OR "Sickle Cell Diseases" [TW])) AND ("Health Services Accessibility"[MeSH] OR "Access to health care" OR "Access to Health Services" OR "Access to Care" OR "Care, Access to" OR "Accessibility of Health Services" OR "Accessibility, Health Services" OR "Availability of Health Services" OR "Health Services Availability" OR "Access To Care, Health" OR "Treatment, Access to" OR "Health Services Geographic Accessibility" OR "Access to Medicines" OR "Access to Medicine" OR "Medicine, Access to" OR "Medicines, Access to" OR "Access to Medications" OR "Access to Medication" OR "Medication, Access to" OR "Medication Access" OR "Access, Medication" OR "Telemedicine" OR "Equitable access" OR "Specialized care" OR "Access to nutrition" OR "Educational Status"[Mesh] OR "Educational Achievement" OR "Status, Educational" OR "Achievement, Educational" OR "Achievements, Educational" OR "Educational Achievements" OR "Educational Status, Maternal" OR "Status, Maternal Educational" OR "Maternal Educational Status" OR "Educational Status, Paternal" OR "Status, Paternal Educational" OR "Paternal Educational Status" OR "Knowledge" OR "Knowledge Base" OR "Higher education" OR "Anticipatory Knowledge" OR "Anticipatory Guidance" OR "Health Literacy" OR "Parental Education" OR "Education of parents" OR "Parental Interest" OR "Caregiver knowledge" OR "Spending power" OR "Affordability of care" OR "Parental protection" OR "parental Income" OR "patient income" OR "Patients adherence" OR "Treatment compliance" OR "Patient follow-up" OR "Continuation of care" OR "Social support" OR "Family support" OR "Financial support" OR "Quality of Life" OR "Health-related quality of life" OR "Socioeconomic factors" [MeSH] OR "Factor, Socioeconomic" OR "Socioeconomic Factor" OR "Factors, Socioeconomic" OR "Standard of Living" OR "Living Standard" OR "Living Standards" OR "Social Inequality" OR "Inequalities, Social" OR "Inequality, Social" OR "Social Inequalities" OR "awareness" OR "interpersonal relationships" OR "social determinants of health" ) | 2903 |
| Limited to 1973 – 2023 and English only |  |  |

## Appendix II: Data extraction instrument

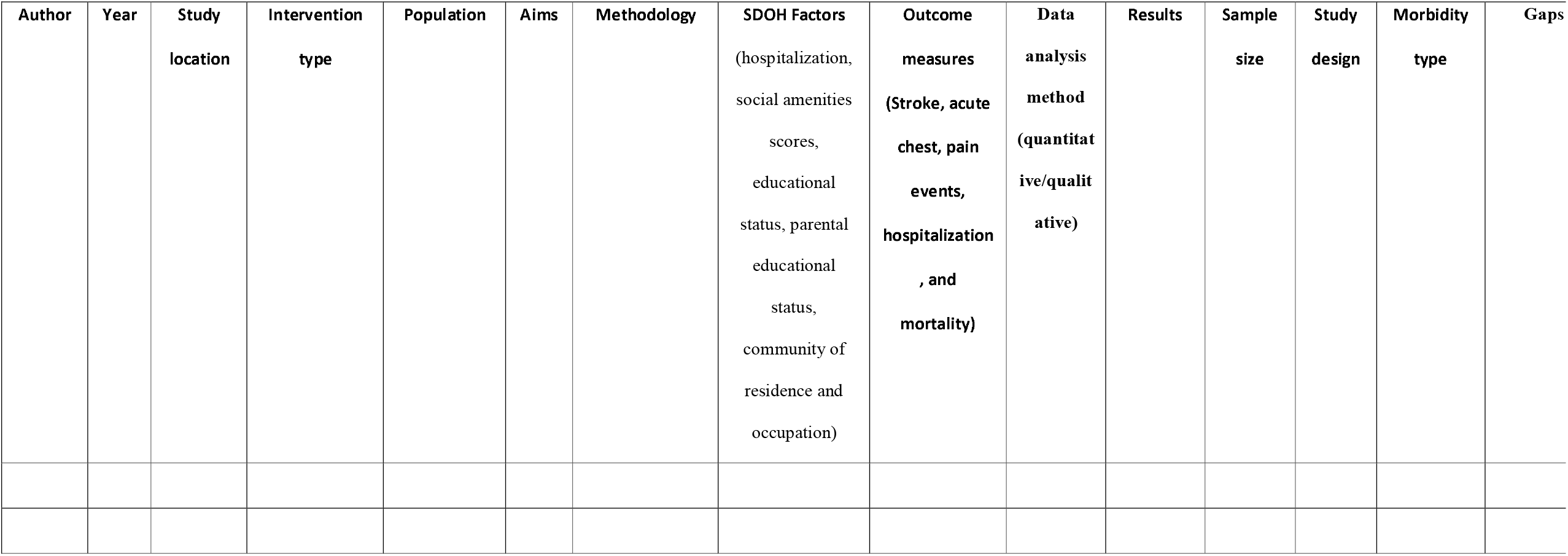
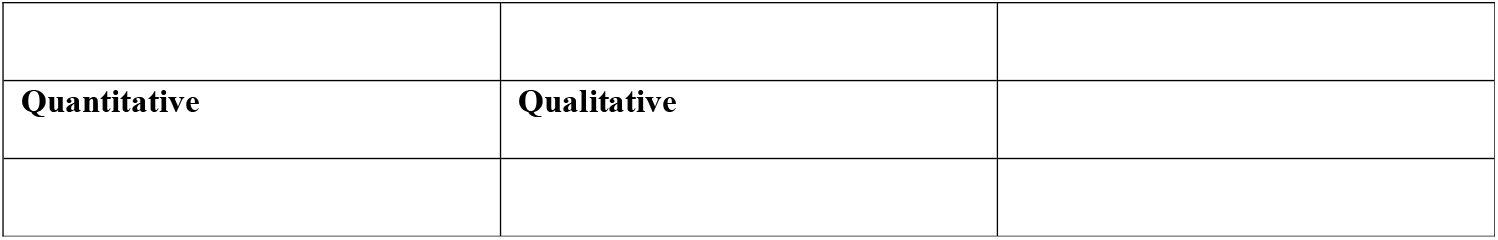

| Author | Year | Study location | Intervention type | Population | Aims | Methodology | SDOH Factors<br>(hospitalization, social amenities scores, educational status, parental educational status, community of residence and occupation) | Outcome measures<br>(Stroke, acute chest, pain events, hospitalization , and mortality) | Data analysis method<br>(quantitative/qualitative) | Results | Sample size | Study design | Morbidity type | Gaps |
| --- | --- | --- | --- | --- | --- | --- | --- | --- | --- | --- | --- | --- | --- | --- |

|  |  |
| --- | --- |
| **Quantitative** | **Qualitative** |

## References

1. Rees DC, Williams TN, Gladwin MT. Sickle-cell disease. Lancet. 2010;376(9757):2018–31. 2.

2. Kato GJ, Piel FB, Reid CD, Gaston MH, Ohene-Frempong K, Krishnamurti L, et al. Sickle cell disease. Nat Rev Dis Primers. 2018;4:18010.

3. Piel FB, Steinberg MH, Rees DC. Sickle Cell Disease. N Engl J Med. 2017;376(16):1561–73.

4. Bhatt K, Reid ME, Lewis NA, Asnani MR. Knowledge and health beliefs of Jamaican adolescents with sickle cell disease. Pediatric Blood & Cancer. 2011;57(6):1044–8.

5. Ware RE, de Montalembert M, Tshilolo L, Abboud MR. Sickle cell disease. Lancet. 2017;390(10091):311–23.

6. Poku BA, Caress A-L, Kirk S. Adolescents’ experiences of living with sickle cell disease: An integrative narrative review of the literature. International Journal of Nursing Studies. 2018;80:20–8. 7.

7. Lubeck D, Agodoa I, Bhakta N, Danese M, Pappu K, Howard R, et al. Estimated Life Expectancy and Income of Patients With Sickle Cell Disease Compared With Those Without Sickle Cell Disease. JAMA Netw Open. 2019;2(11):e1915374.

8. Wilkinson R, Marmot M, World Health Organization. Regional Office for E. Social determinants of health: the solid facts. 2nd (en) ed. Copenhagen: World Health Organization. Regional Office for Europe; 2003 2003.

9. Addressing Social Determinants to Improve Patient Care and Promote Health Equity: An American College of Physicians Position Paper. Annals of Internal Medicine. 2018;168(8):577–8.

10. Berghs M, Ola B, Cronin De Chavez A, Ebenso B. Time to apply a social determinants of health lens to addressing sickle cell disorders in sub-Saharan Africa. BMJ Global Health. 2020;5(7):e002601. 11.

11. Morton S, Pencheon D, Squires N. Sustainable Development Goals (SDGs), and their implementation: A national global framework for health, development and equity needs a systems approach at every level. British medical bulletin. 2017;124:1–10.

12. contributors P. Biopsychosocial Model: Physiopedia; [updated 7 November 2022 23:13 UTC. Available from: https://www.physio-pedia.com/index.php?title=Biopsychosocial_Model&oldid=319922.

13. Novelli EM, Gladwin MT. Crises in Sickle Cell Disease. Chest. 2016;149(4):1082–93.

14. Alishlash AS, Rutland SB, Friedman AJ, Hampton JI, Nourani A, Lebensburger J, et al. Acute chest syndrome in pediatric sickle cell disease: Associations with racial composition and neighborhood deprivation. Pediatr Blood Cancer. 2021;68(4):e28877.

15. Serjeant GR, Serjeant BE. Sickle Cell Disease. Third ed. The United States: Oxford University Press; 1985 2001. 772 p.

16. Granja PD, Quintão SBM, Perondi F, de Lima RBF, Martins CLM, Marques MA, et al. Leg ulcers in sickle cell disease patients. J Vasc Bras. 2020;19:e20200054.

17. Altman IA, Kleinfelder RE, Quigley JG, Ennis WJ, Minniti CP. A treatment algorithm to identify therapeutic approaches for leg ulcers in patients with sickle cell disease. Int Wound J. 2016;13(6):1315–24.

18. Serjeant GR, Serjeant BE, Mohan JS, Clare A. Leg ulceration in sickle cell disease: medieval medicine in a modern world. Hematol Oncol Clin North Am. 2005;19(5):943–56, viii-ix.

19. Minniti CP, Kato GJ. Critical Reviews: How we treat sickle cell patients with leg ulcers. Am J Hematol. 2016;91(1):22–30.

20. Cintron-Garcia J, Ajebo G, Kota V, Guddati AK. Mortality trends in sickle cell patients. Am J Blood Res. 2020;10(5):190–7.

21. Onimoe G, Rotz S. Sickle cell disease: A primary care update. Cleveland Clinic Journal of Medicine. 2020;87(1):19.

22. Serjeant GR, Chin N, Asnani MR, Serjeant BE, Mason KP, Hambleton IR, et al. Causes of death and early life determinants of survival in homozygous sickle cell disease: The Jamaican cohort study from birth. PLoS One. 2018;13(3):e0192710.

23. Pollitt RA, Rose KM, Kaufman JS. Evaluating the evidence for models of life course socioeconomic factors and cardiovascular outcomes: a systematic review. BMC Public Health. 2005;5(1):7.

24. Khan H, Krull M, Hankins J, Wang W, Porter J. Sickle Cell Disease and Social Determinants of Health -- A Scoping Review. Authorea Preprints; 2022.

25. Page MJ, McKenzie JE, Bossuyt PM, Boutron I, Hoffmann TC, Mulrow CD, et al. The PRISMA 2020 statement: an updated guideline for reporting systematic reviews. BMJ. 2021;372:71.

26. Moher D, Shamseer L, Clarke M, Ghersi D, Liberati A, Petticrew M, et al. Preferred reporting items for systematic review and meta-analysis protocols (PRISMA-P) 2015 statement. Syst Rev. 2015;4(1):1.

27. Peters M, Godfrey C, McInerney P, Munn Z, Trico A, Khalil H. Chapter 11: Scoping Reviews. 2020.

28. Westphaln KK, Regoeczi W, Masotya M, Vazquez-Westphaln B, Lounsbury K, McDavid L, et al. From Arksey and O’Malley and Beyond: Customizations to enhance a team-based, mixed approach to scoping review methodology. MethodsX. 2021;8:101375.

29. Buus N, Nygaard L, Berring LL, Hybholt L, Kamionka SL, Rossen CB, et al. Arksey and O’Malley’s consultation exercise in scoping reviews: A critical review. J Adv Nurs. 2022;78(8):2304–12.

